# Cross-modal sensory boosting to improve high-frequency hearing loss

**DOI:** 10.1101/2023.06.01.23290351

**Authors:** Izzy Kohler, Michael V. Perrotta, Tiago Ferreira, David M. Eagleman

## Abstract

High frequency hearing loss is one of the most common problems in the aging population and with those who have a history of exposure to loud noises. To address high-frequency hearing loss, we developed a multi-motor wristband that uses machine learning to listen for specific high frequency phonemes. The wristband vibrates in spatially unique locations to represent which phoneme was present, in real time. We recruited 16 participants with high frequency hearing loss and asked them to wear the wristband for six weeks. Their degree of disability associated with hearing loss was measured weekly using the Abbreviated Profile of Hearing Aid Benefit (APHAB). Our findings show that vibrotactile sensory substitution delivered by a wristband that produces spatially distinguishable vibrations in correspondence with high frequency phonemes helps individuals with high frequency hearing loss improve their understanding of verbal communication. We find that vibrotactile feedback provides benefits whether or not a person wears hearing aids, albeit in slightly different ways. Finally, our results also demonstrate that individuals with the greatest difficulty understanding speech prior to the study experience the greatest amount of benefit from vibrotactile feedback.

## Introduction

Hearing loss affects 466 million people worldwide (Olusanya et al., 2019). High frequency hearing loss is one of the most common types of hearing loss and renders high-pitched sounds, such as the voices of women and children, more difficult to hear (Chang et al., 2011). It can affect people of any age, but is more common among older adults and people who have been repeatedly exposed to loud noises (Michels et al., 2019). This type of hearing loss can be frustrating and disabling, making it difficult to understand speech communication and interact effectively with the world, leading to a decline in quality of life and isolation (Michels et al., 2019).

Individuals with high-frequency hearing loss struggle to hear consonants with higher frequency sound components, such as *s, t, and f*. As a result of the hearing loss, speech is reported as sounding muffled, most noticeably in noisy environments. Commonly, people with age related hearing loss (presbycusis) will report that they can hear but cannot understand. It is often noticed when a person has trouble understanding women’s and children’s voices, as well as with detecting other sounds such as the ringing of a cell phone or the chirping of birds. Assistive hearing technologies such as hearing aids (HA) and cochlear implants (CI) are able to offer some assistance with understanding speech communication, however they have limitations: One of the most commonly reported disappointments among users of HAs and CIs is that they still cannot understand speech (Hickson et al., 2014).

The auditory cortex is activated by vibrotactile information in individuals who are hearing impaired and deaf (Auer et al., 2007; Levänen et al., 1998). Auditory and vibratory stimuli are also similar in temporal patterning. This makes tactile vibrations an ideal modality for cross-modal plasticity of auditory input (Soto-Faraco & Deco, 2009). Levänen et al. (1998) used recorded Magnetoencephalography (MEG) signals to demonstrate activation of the supratemporal auditory cortex when vibrotactile stimuli was applied to the palm and fingers of a congenitally deaf adult. Additionally, the activity pattern within the auditory cortices indicated discrimination between 180 Hz frequency vibrations and 250 Hz frequency vibrations in the auditory cortex that was similar to the discrimination patterns for similar sound frequencies in normal hearing adults. In a similar experiment, Auer et al. (2007) used fMRI imaging to show that vibration stimuli derived from speech and administered to the hands of adults with early onset deafness and long-term hearing aid use recruited widespread activity within their auditory cortical regions. Auditory stimulation and vibrotactile stimulations share common characteristics, stimulate mechanoreceptors during the process of cognitive interpretation, and can be naturally perceived as an integrated signal processed in common brain regions (Soto-Faraco & Deco, 2009).

To address the speech understanding limitations associated with high-frequency hearing loss, we have developed a vibrotactile sensory substitution solution in the form of a wristband (Novich & Eagleman, 2015; Perrotta et al., 2021; Eagleman & Perrotta, 2023). This device delivers spatially unique vibrations to the wrist in correspondence with target phonemes that are commonly difficult for individuals with presbycusis to detect. The wristband receives sound from the environment through an onboard microphone and uses a machine learning algorithm to filter background noise and extract target phonemes from speech. Each phoneme signal is mapped to its own unique linear resonant actuator (LRA) in the strap of the wristband where it is felt as a vibration on the skin. There are four LRAs embedded within the wristband strap, giving each target phoneme a unique spatial location on the wrist. Parts of speech that are audible to the user are unconsciously integrated with the spatially unique vibratory signals representing the inaudible portions of speech. The user is then able to understand a complete and meaningful message through integration of the complementary sensory inputs.

Our own prior work in this area demonstrated that when two words are algorithmically translated into spatiotemporal patterns of vibration on the skin of the wrist, they are distinguishable to individuals who are hard of hearing or deaf up to 83% of the time for two words that are similar and up to 100% of the time for two words that are not similar (Perrotta et al., 2021; Cieśla et al., 2022). Further studies showed that sound-to-touch sensory substitution devices may help people with hearing impairments, allowing them to access sensory information otherwise unable to be accessed. Weisenberger & Russell (1989) used single channel vibrotactile aids designed to translate acoustic stimuli into representative vibration patterns on the wrist to improve performance on environmental sound identification tests from 55% to 95% correct and improve performance on single word identification testing from 60% to 90%.

In this study, we demonstrate that a simple wearable sensory substitution device that transforms speech sounds into haptic vibrations on the wrist can help individuals with high frequency hearing loss better understand speech communication throughout their normal daily routine.

This will improve the quality and productivity of their daily interactions, allow them to enjoy audio based entertainment such as movies and podcasts, help them understand conversations in complicated acoustic environments, and fill the residual gaps of impairment left by their hearing aids.

## Methods

### Device

Participants wore a haptic wristband (**Figure 1**) that vibrates to indicate the occurrence of specific phonemes. The wristband contains four vibrating motors embedded in the wrist strap, a microphone, a power button, a microcontroller, and a battery.

**Figure 1.**
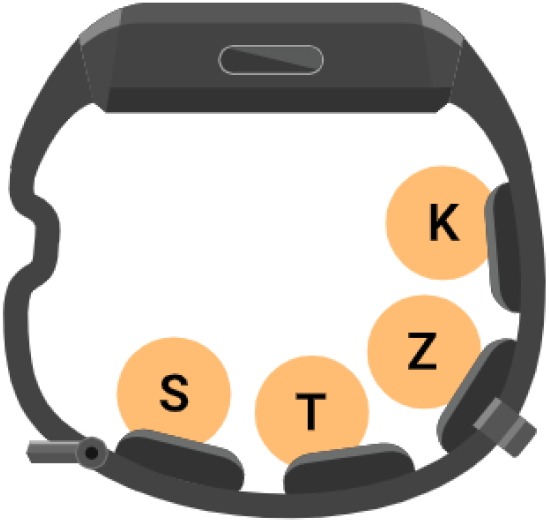
The Neosensory wristband has four vibrating motors embedded in the wrist strap. The top of the wristband contains a power button and a microphone. Each phoneme is assigned to an independent motor.

The motors are linear resonant actuators (LRAs) which vibrate in a sine wave and are capable of rising from 0 to 50% of their maximum amplitude within 30 ms. The motors vibrate at 175 Hz, the frequency at which human skin has the highest sensitivity (Verrillo, 1980). Each motor vibrates at 1.7 GRMS (16.6 m/s^2^). The motors are separated from one another at a distance of 18.2 mm and 19.2 mm for the small and large wristband sizes, respectively (center-to-center distances). Each motor pad contacts the wearer’s skin on a rectangular area that measures approximately 8.2 mm by 8.5 mm.

The top of the wristband is a module that contains the power button, a microphone, and a microcontroller. The microphone captures audio and sends this data to the microcontroller. The microcontroller processes the audio data through a phoneme-detection algorithm and vibrates the motors according to the output of the algorithm.

### Algorithm

The algorithm processes incoming audio to determine when any target phoneme is detected. If a target phoneme is detected, the corresponding motor vibrates for 80 ms.

The four target phonemes were /s/, /t/, /z/, and /k/. Each motor on the wristband was assigned to a different target phoneme. Figure 1 shows the motor assignments for each phoneme. In addition to the real algorithm, we also created a sham algorithm for testing purposes. The real algorithm captures audio through a microphone and inputs this audio into a machine learning algorithm, which detects when one of the target phonemes occurred. The sham algorithm detects when speech is occurring and causes the motors to vibrate at the same occurrence rate as the real algorithm, but without correspondence to any specific speech sound. For instance, if the real algorithm detects an /s/ on average 4 times during 1 minute of speech, the sham algorithm will cause the /f/ motor to vibrate at a rate of 4 times per minute while speech is detected, but at random without any correlation to the detection of /f/ phonemes.

The four phonemes were chosen based on a combination of the following three factors: (1) how difficult each phoneme is for hearing impaired listeners to hear, (2) how frequently each phoneme occurs in spoken English, and (3) how well our algorithm can detect each phoneme. The difficulty was pooled from several studies of phoneme confusion for hearing impaired listeners. (Phatak et al., 2009) asked older hearing impaired listeners to identify the consonant in a presented consonant-vowel (CV) syllable. (Woods et al., 2015) presented the California Syllable Test, which uses consonant-vowel-consonant (CVC) syllables, to older hearing impaired listeners in both aided (with hearing aids) and unaided conditions. (Sher & Owens, 1974) presented a four-alternative forced choice test with CVC syllables, where only either the initial or final consonant differed between choices. Synthesizing the results of these three studies, we found that the following consonants are the most difficult to hear for a listener with presbycusis: /dh/, /th/, /ng/, /v/, /b/, /hh/, /f/, /z/, /s/, /t/. Of these, /th/ and /ng/ are present in spoken English less than 1% of the time (Mines et al., 1978). Our algorithm performed poorly on /dh/, /b/, /f/, and /hh/.

### Phoneme Detection

The algorithm consists of feature extraction and inference engine components. The feature extraction module segments an audio stream captured from the microphone into 32 ms frames with 16 ms of overlap. Each audio frame undergoes analysis to extract distinct features suitable for phoneme recognition. The features are also subject to further processing which amplifies phoneme-specific information they contain and ensures robustness towards continuously changing environmental conditions.

The inference engine takes these feature vectors and outputs phoneme predictions. The core of the inference engine is a neural network model which uses a Real Time Temporal Convolutional Network (RTTCN) structure optimized for real-time speech recognition. The full latency from phoneme onset to vibration onset is 170 ms.

### Paradigm

Participants wore the wristband every day for six weeks. Each day the participants were required to spend at least one hour watching television or listening to an audiobook, podcast, or other speech-based media while wearing the wristband. The instructions were to choose something engaging so their attention would be directed toward understanding what was being said, while the wristband provided the assistive haptic feedback. The purpose of this required daily exercise was to ensure the participant was immersed in a minimum amount of active listening each day so the brain would learn to integrate the audible speech sounds with the haptic vibratory representations of the inaudible speech sounds to form a complete meaning. In addition to the required hour of practice, participants were encouraged to wear the wristband whenever engaged in conversation or active listening to speech communication.

### Tasks

#### APHAB

Prior to starting the study, and at the end of each week during the study, participants completed a modified version of the Abbreviated Profile of Hearing Aid Benefit (APHAB), in which we removed the six questions included in the aversiveness subscale (Cox, 1997). These questions were removed because they are asking about the unpleasantness of sounds heard through a hearing aid, which does not apply for our device. The remaining 18 questions on the APHAB ask questions about one’s ability to understand verbal communication in different scenarios.

For example, one of the questions is “When I am in a crowded grocery store, talking with the cashier, I can follow the conversation.” In the conventional questionnaire, participants answer the questions independently for their experiences while using and while not using their hearing aids. In the current study, participants answered the questions independently for their experiences while using and while not using the wristband. If the participant regularly wore hearing aids, “with the wristband” was referring to wearing the wristband in addition to their hearing aids and “without the wristband” was referring to wearing their hearing aids alone. The benefit score is calculated by subtracting the final aided score at the conclusion of the trial from the baseline unaided score that was measured at the beginning of the trial.

#### Final Questionnaire

On the final day of the study, participants answered a questionnaire which asked two questions, both on a Likert scale from 1 to 10: “How much did the Clarify wristband help you understand speech?” and “How likely are you to recommend the Clarify wristband to a friend or colleague?”.

#### Participants

We recruited participants via online advertising for a paid study related to hearing loss. Participants were eligible if they were between 18 and 80 years of age, had access to a mobile (i.e., iOS or Android) device and a computer, primarily spoke English, and met the following criteria for high frequency hearing loss: a pure-tone audiogram, either produced by an audiologist in the past 24 months or from an audiogram mobile app (e.g. Mimi: https://mimi.health/hearing-test-apps), must show at least 55 dB of hearing loss at 4 kHz averaged across both ears (with neither ear’s 4 kHz threshold being less than 40 dB HL) and no more than 35 dB HL averaged across both ears and across 500 Hz and 1000 Hz tones. Sixteen eligible participants completed the study. The average age was 68.8 (11.6) years. The type and severity of hearing loss was determined from pure tone audiograms. The average pure tone threshold of both ears at 500 Hz and 1000 Hz was 30 (13) dB and the average pure tone threshold of both ears at 4000 Hz was 63 (9) dB (**Figure 2**).

**Figure 2.**
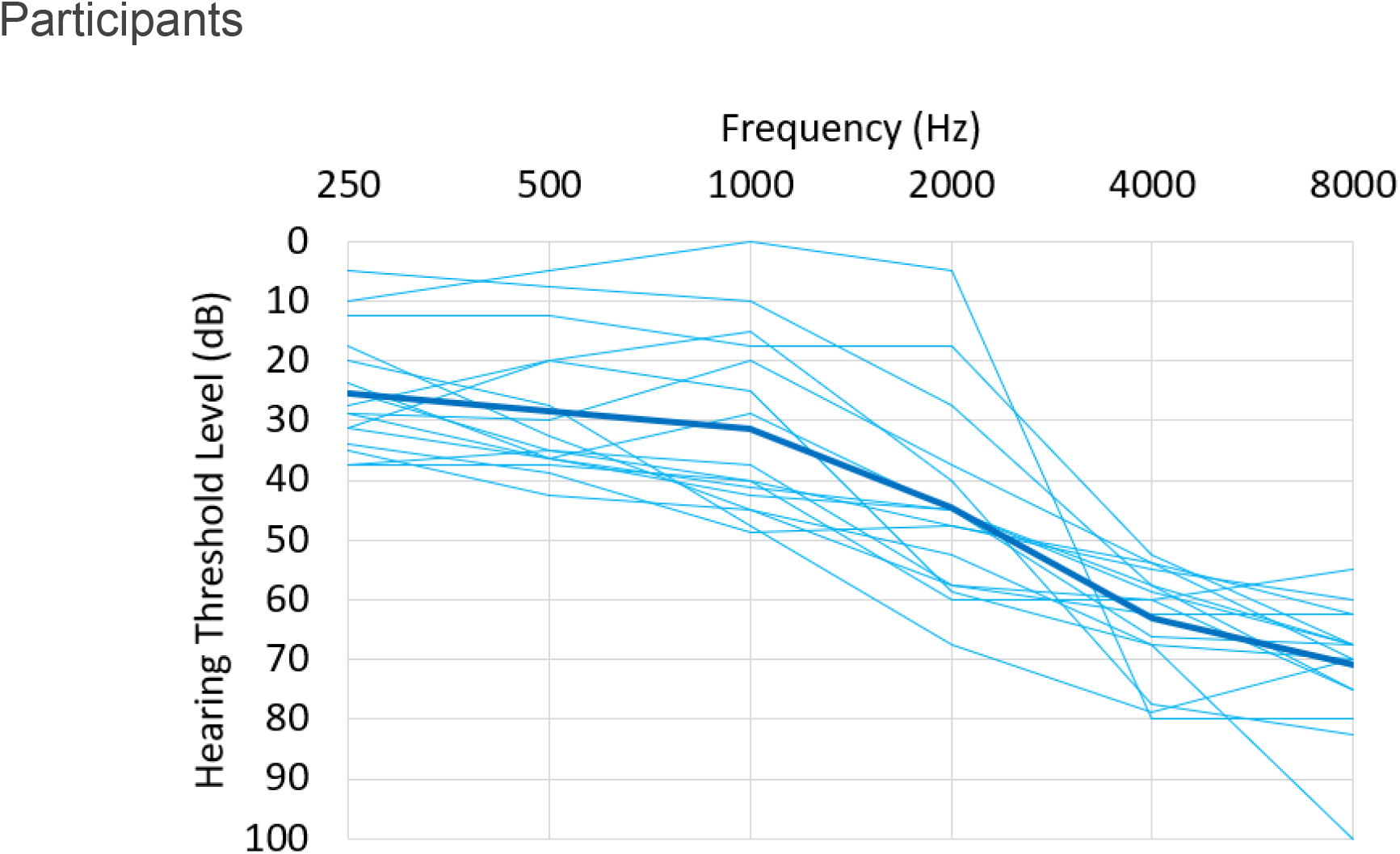
The average pure tone audiogram of both ears. The thin lines represent each individual participant, the thick line represents the group average

## Results

As shown in **Figure 3**, after only one week of wearing the wristband daily, the average APHAB benefit score (*unaided - aided*) was 8.61 points, with a baseline score of 40.32 points that dropped to 31.71 points (SD=12.11, n=16, p=.01, two-tailed dependent t-test). Baseline is defined as the unaided APHAB score taken prior to starting to use the wristband. As a reminder, if the participant regularly used hearing aids, they were asked to answer the unaided questions based on how they felt with their hearing aids on. If the participant never used hearing aids, they were asked to answer the unaided questions based on how they felt without any hearing assistance. The average aided APHAB score continued to drop at a slower, more steady pace for the remaining five weeks of the study. By the end of the six week study, the average APHAB benefit score had reached a clinically meaningful and statistically significant value of 12.39 points (Cox & Alexander, 1995) from a baseline of 40.32 to a final score of 27.93 (SD=13.11, n=16, p=.002, two-tailed dependent t-test).

**Figure 3.**
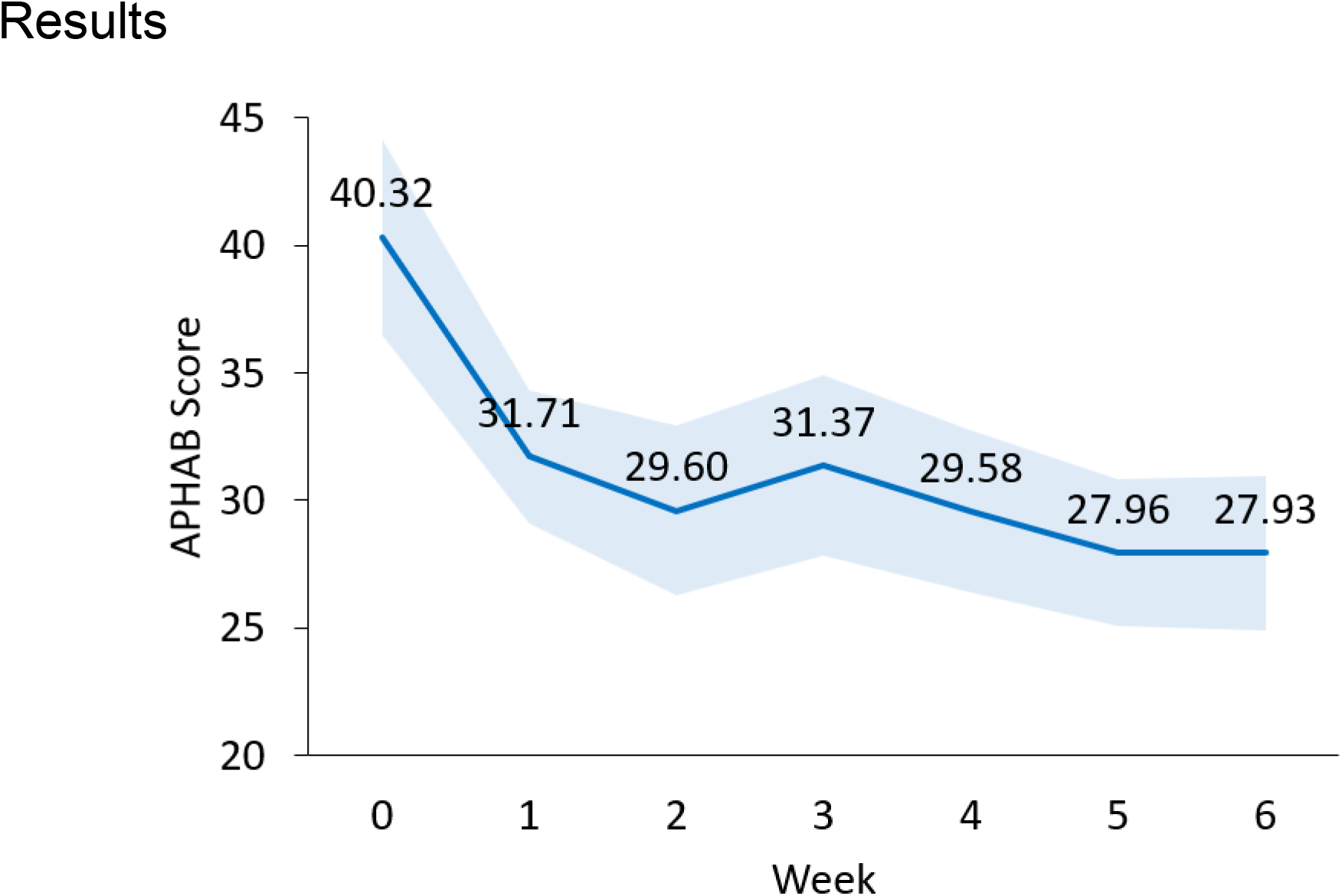
Six week progression of the Abbreviated Profile of Hearing Aid Benefit (APHAB) scores. Error boundary represents standard error of the mean. Week 0 score is the unaided APHAB score (prior to starting with the wristband); subsequent weeks show the aided APHAB score (with the wristband).

Simple linear regression analysis was used to test if a participant’s baseline APHAB score explains their benefit over baseline APHAB score after the six weeks (**Figure 4**). The results of the regression indicate that the average baseline score explains 43% of the variation in the average APHAB benefit at six weeks (F(1,14)=10.55, p=.006). These results are significant at the p<.05 level.

**Figure 4.**
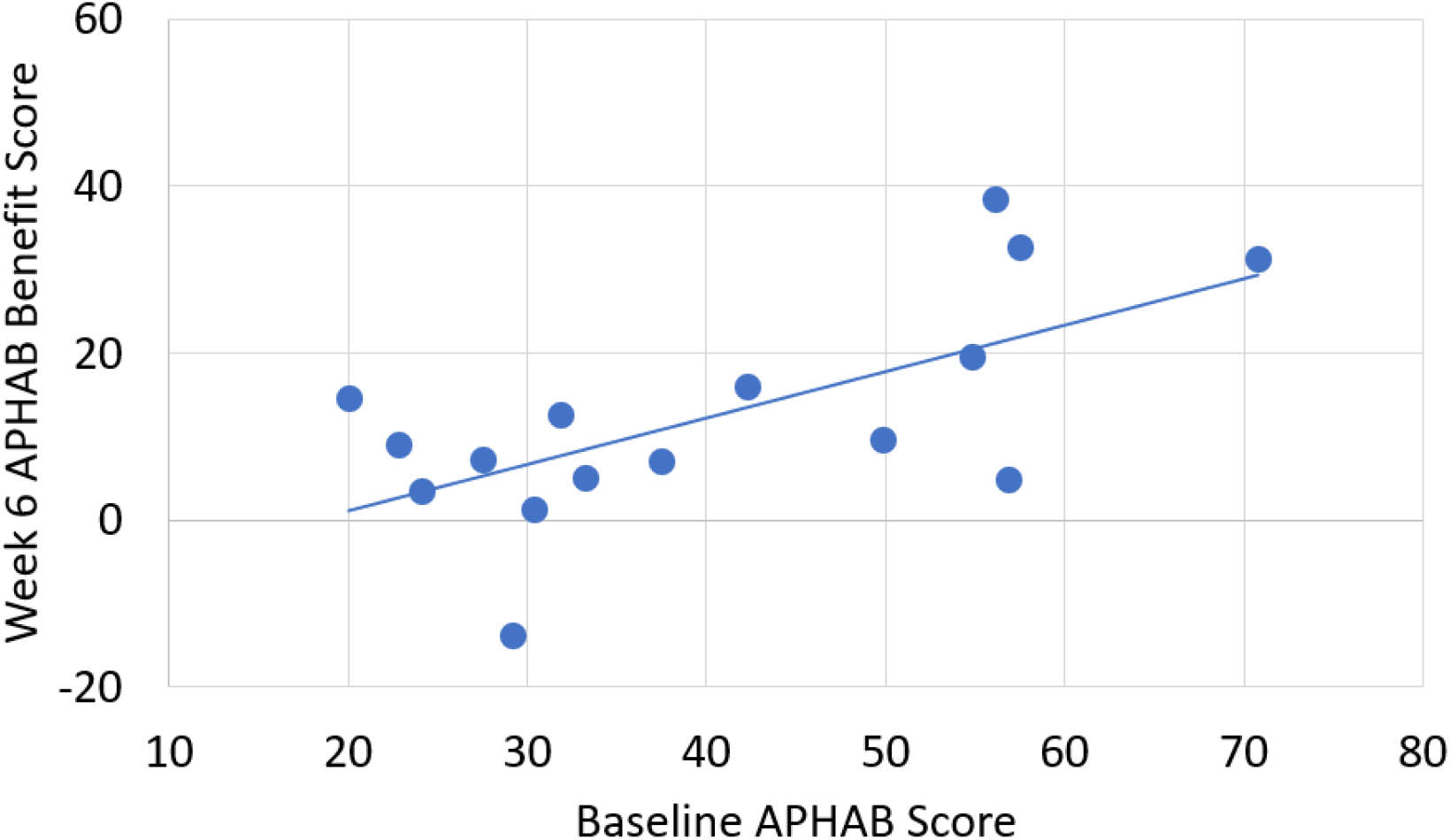
Scatter plot of the baseline APHAB score vs the final APHAB benefit. The linear regression line demonstrates the correlation between the degree of disability without the assistance of Clarify at baseline and the final benefit score at week 6 with the aid of Clarify.

We compared participants who used hearing aids to those who did not. Nine of the participants used hearing aids to help them understand speech and seven of the participants did not.

Results showed a 10.78 point difference between the two subgroups in the average APHAB benefit score at six weeks (t(14)=2.14, p=.10, two-tailed independent t-test, **Figure 5**). While the difference in benefit score between the two subgroups was not statistically significant, it did reach the ten point threshold for clinical relevance (Cox, 1997; Cox & Alexander, 1995). Also, while the subgroup without hearing aids started the study at a higher level of disability, they ended the study at a lower level of disability. The subgroup without hearing aids started with a baseline APHAB score of 44.09 (16.66) points while the subgroup with hearing aids started with a baseline score of 37.40 (14.61) points. The subgroup without hearing aids concluded the study with an APHAB score of 25.63 (12.51) points while the subgroup with hearing aids concluded the study with an APHAB score of 29.72 (12.01) points. Another noteworthy difference between the subgroups was the group that did not wear hearing aids demonstrated both a statistically significant and clinically meaningful aided APHAB benefit from baseline, while the subgroup that did wear hearing aids did not. The subgroup that did not wear hearing aids ended the study with an average APHAB benefit over baseline of 18.45 points (SD=11.70, n=7, p=.005, two-tailed dependent t-test). The subgroup that wore hearing aids ended the study with an average APHAB benefit over baseline of 7.67 points (SD=12.730, n=9, p=.108, two-tailed dependent t-test)

**Figure 5.**
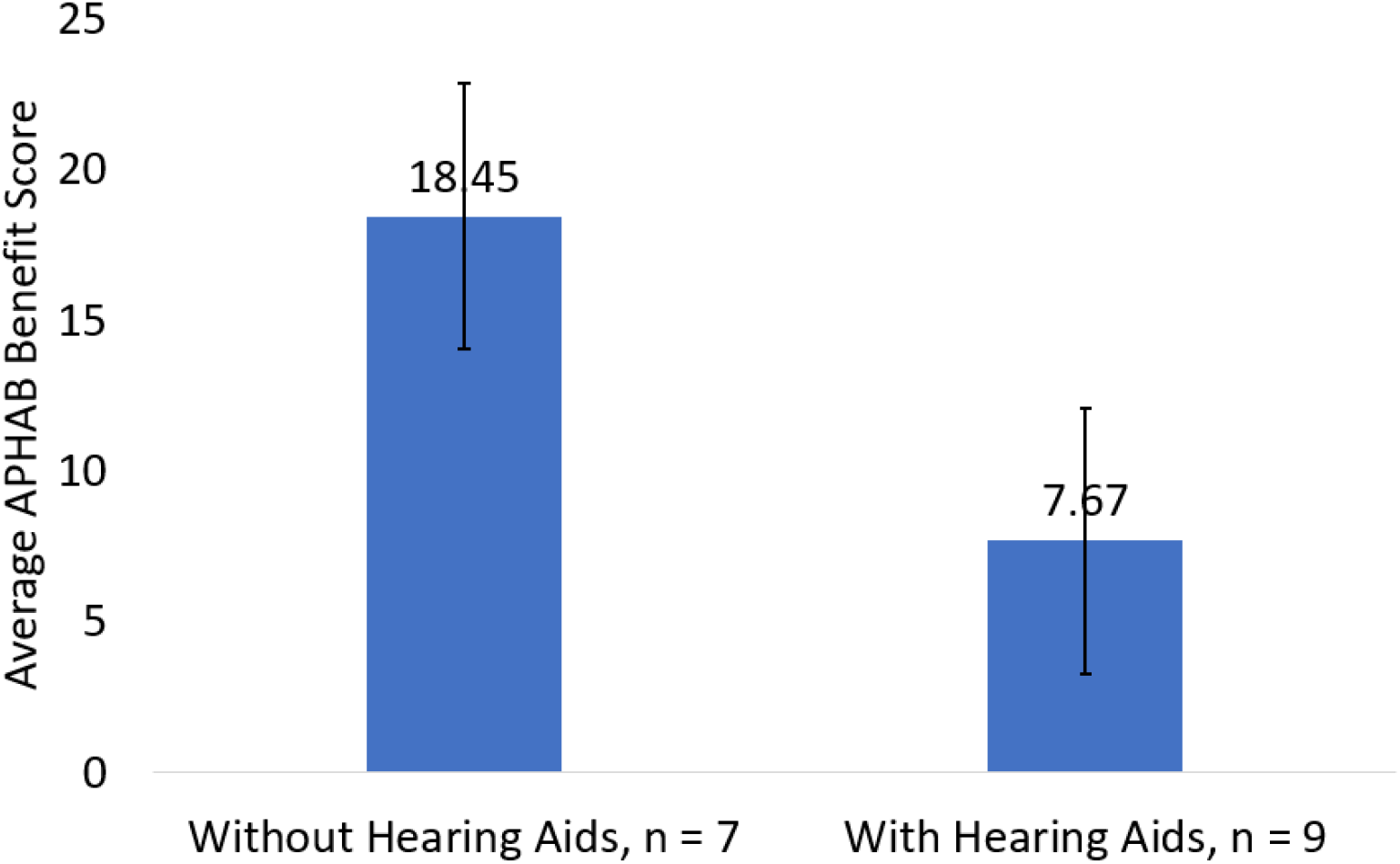
On average, the subgroup of participants who were not users of hearing aids ended the study with a higher benefit score than the subgroup of participants who were regular users of hearing aids. Error bars represent standard error of the mean.

Subscale analyses were performed for ease of communication (EOC), background noise (BN), and reverberation (RV) (**Figure 6**). These subscales are reflective of speech communication under ideal conditions, in noisy environments, and in reverberant environments (Cox & Alexander, 1995). The average benefit score above baseline for EOC was 15.44 (SD=13.88, n=16, p<.001, two-tailed dependent t-test). Those who wore hearing aids and those who did not wear hearing aids had similar EOC benefit scores: 13.57 (SD=15.71, n=9, p=.03, two-tailed dependent t-test) and 17.83 (SD=11.85, n=7, p=.007, two-tailed dependent t-test) respectively. The average benefit score above baseline for BN was 10.88 (SD=17.54, n=16, p=.03, two-tailed dependent t-test), with a 16.99 point difference in BN benefit between those who wore and did not wear hearing aids (no hearing aids 20.43 benefit, hearing aids 3.44 benefit, t(14)=2.14, p=.05, two-tailed independent t-test). The average benefit score above baseline for RV was 10.84 (SD=16.95, n=16, p=.02, two-tailed dependent t-test), with a 11.12 point difference in RV benefit between hearing aids and no hearing aids (HA=17.10, no HA=5.98, t(14)=2.14, p=.20, two-tailed independent t-test).

**Figure 6.**
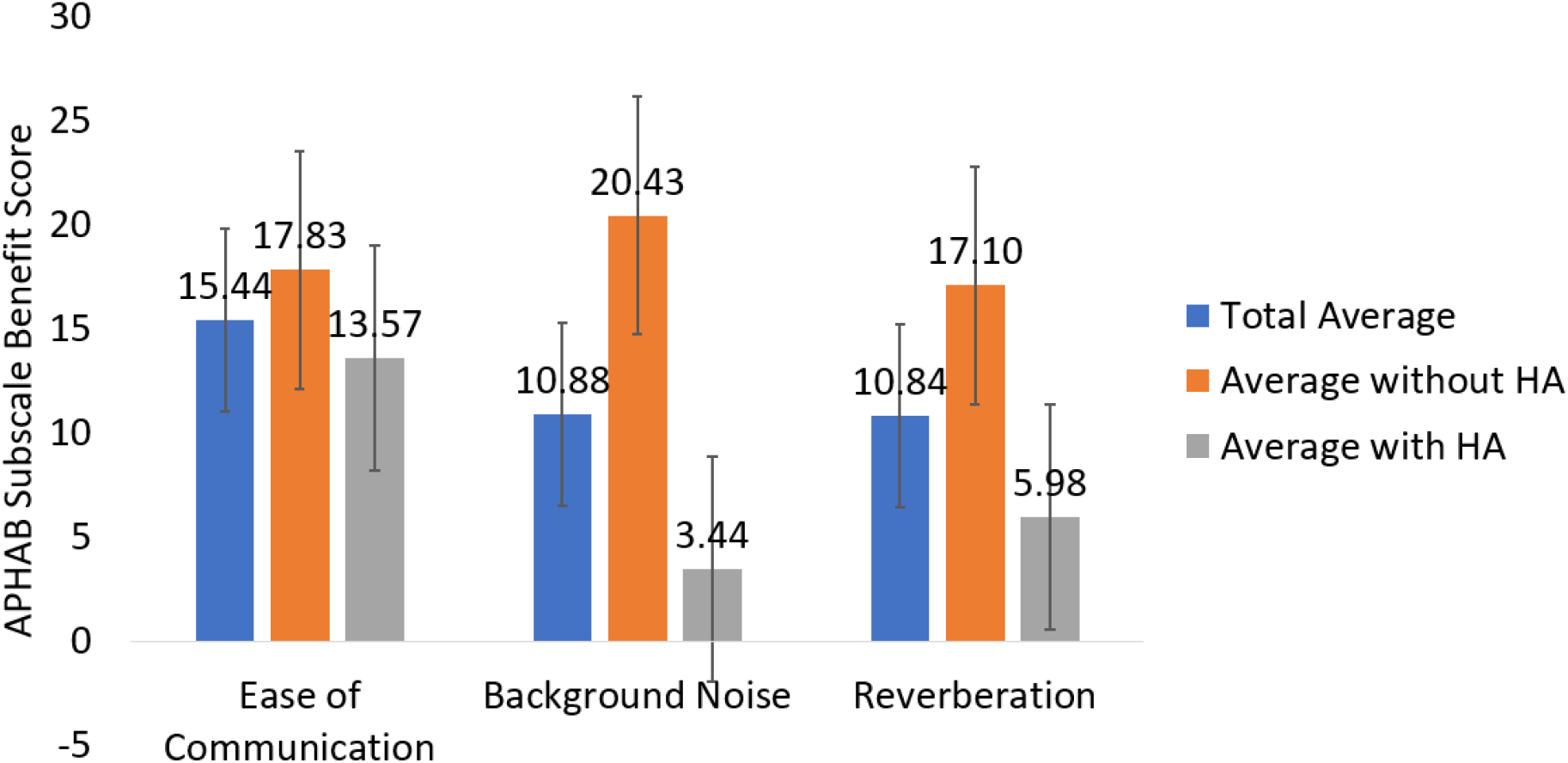
Average APHAB subscale benefit scores for the entire participant group, subgroup who were regular users of hearing aids, and the subgroup that did not wear hearing aids. There were 16 participants total, 9 who were regular users of hearing aids, and 7 who did not use hearing aids. Error bars represent standard error of the mean.

## Discussion

In this study we expanded on our prior work which showed deaf and hard of hearing individuals are capable of identifying sound categories through patterns of vibration applied to the wrist (Perrotta et al., 2021). Here, we demonstrated that individuals with high frequency hearing loss are able to improve their understanding of speech communication using vibrational representations of high frequency speech sounds on the wrist. Results demonstrate that after one week of wearing the wristband, participants were able to improve their ability to understand conversations during daily interactions. They then continued to improve over the course of the 6-week study, at a slower rate. This reflects prior research findings of an innate ability for those with hearing loss to rapidly learn to interpret tactile vibrations as a substitute for audio information (Soto-Faraco & Deco, 2009). The understanding of vibrations is further strengthened and perfected over time with practice as the portions of the auditory cortex that respond to tactile vibration expands (Auer et al., 2007; Good et al., 2014; Levänen et al., 1998). We further found that participants who started the study with a higher APHAB score experienced a greater improvement in their ability to understand speech by the end of the six week trial. Out of 16 participants, 14 ended the study with an APHAB score of 40 or below (which translates to difficulty understanding speech less than half of the time). Five participants started the study with an unaided APHAB score of 50 points or higher; for three of them, the final APHAB benefit score was >30 points. One potential hypothesis for why participants who started the trial with greater difficulty understanding speech experience greater improvement is that more of their auditory cortex is available for the interpretation of tactile sound representation (Auer et al., 2007). This could be an interesting topic for future research.

Participants without hearing aids benefitted the most from vibrotactile sensory substitution for speech understanding. Given that this group started the study with a higher APHAB score (above), we presume the difference is because the hearing aids group already gains benefit from their technology and therefore has less room for improvement. It is difficult to predict what the interaction between hearing aids and vibrotactile feedback will be because of the differing signal processing techniques used in digital hearing aid technologies. Digital hearing aids convert sound waves into numerical codes before amplifying them. This code contains information about a sound’s frequency and amplitude, allowing the hearing aid to be specially programmed to amplify some frequencies more than others. Digital sound processing capabilities allow an audiologist to adjust the hearing aid to a user’s needs and to different listening environments. Digital hearing aids can also be programmed to focus on sounds coming from a specific direction. It is possible the wristband represents sounds that differ significantly from those represented by the hearing aid. Future studies can possibly explore directly connecting the wristband to the user’s hearing aids through a bluetooth signal so that the wristband’s signals directly correspond with the sounds the user is hearing.

Individuals with hearing impairment have great difficulty understanding speech in the presence of background noise. It is one of the primary complaints expressed by many with hearing loss, and one of the most difficult impairments to resolve. Individuals with hearing loss are unable to resolve the closely spaced harmonics of speech sounds to perform a spectral analysis with enough detail to extract the time-frequency portions of the speech that are relatively spared from corruption by the noise background (McArdle & Wilson, 2009). The background noise modulators in hearing aids have not been shown highly effective at helping in these situations (Healy & Yoho, 2016). In this study, we demonstrated the addition of vibrotactile feedback in the presence of background noise enabled individuals who did not wear hearing aids to hear speech communication better (**Figure 5**). Interestingly, the final average BN score for the subgroup without hearing aids was 28.95 (16.15, n=7) and the final average BN score for the subgroup with hearing aids was 40.04 (18.78, n=9) suggesting that those who use hearing aids may benefit from using vibrotactile feedback during conversations in background noise instead of using their hearing aids.

Reverberation is the persistence of a sound after it is produced and is created when the sound is reflected off of surfaces or objects. It is most noticeable when the source of the sound has stopped, but the reflections continue. As the sound reflects off of surfaces and is absorbed by others, the quality of the sound degrades. Every room or outdoor environment has a different level of reverberation due to the construct of the room or area, the reflectiveness of the materials, and the objects in it. Reverberation is natural to every area, but in areas where the reverberation is very high, it can reduce speech intelligibility, especially when background noise is also present. Individuals with hearing loss, including users of hearing aids, frequently report difficulty in understanding speech in reverberant, noisy situations (Cueille et al., 2022). Most hearing aids, both digital and analogue, have limited ability to help individuals with hearing loss in areas of high reverberation (Reinhart et al., 2016). Similar to our findings in background noise, we also found that addition of vibrotactile haptic vibration to the wrist in reverberant environments helped the participants without hearing aids the greatest (**Figure 6**). At the end of the trial, the group of participants who did not wear hearing aids showed an average RV score that was less than the average for the group who were regular hearing aid users. It is possible that individuals who use hearing aids may find haptic vibrations to be more helpful in reverberant environments when the hearing aids are removed because it would eliminate any conflict between the digital processing of the hearing aid and the vibrational signals that are providing information about the sounds of speech without processing.

Ease of communication in the context of the APHAB describes the effort involved in communication under relatively easy listening environments. The interesting discovery from our results was that individuals who use hearing aids experienced a significant improvement in their understanding of conversations under easy listening conditions. In easy listening environments where hearing aids help the most, and perform the least amount of digital signal processing, the addition of haptic vibrations added the greatest amount of additional benefit. Upon completion of the trial, the average EOC score for the subset of participants who were users of hearing aids was 14.65 (SD=6.99, n=9) indicating little to no difficulty understanding speech in easy listening environments. For the subset of participants who were not users of hearing aids, the average EOC score upon completion of the trial was 16.88 (7.73, n=7). Even without the additional help of hearing aids, these participants ended the study with an equivalent capability for understanding speech in easier listening environments, despite starting the trial with a higher level of disability (**Figure 6**).

## Conclusion

We have demonstrated that vibrotactile sensory substitution helps individuals with high frequency hearing loss improve their understanding of verbal communication. The device demonstrated here is a wristband that delivers spatially distinguishable vibrations to the wrist in correspondence with high frequency phonemes. We found that vibrotactile feedback provides more benefit for those without hearing aids than for those with hearing aids, although it does provide benefit for both. This may be a result of how the digital signal processing in hearing aids alters sound for the user in environments with a lot of background noise or reverberation.

Finally, our results also demonstrated that individuals who had the greatest amount of difficulty understanding speech prior to starting the trial experienced the greatest amount of benefit from vibrotactile feedback. Future studies will focus on quantifying the maximum benefits possible and how long improvements continue before a plateau is reached.

## Data Availability

All data produced in the present study are available upon reasonable request to the authors.

